# Spinocerebellar ataxia type 2 has multiple ancestral origins

**DOI:** 10.1101/2023.09.12.23295432

**Authors:** Lucas Schenatto Sena, Gabriel Vasata Furtado, José Luiz Pedroso, Orlando Barsottini, Mario Cornejo-Olivas, Paulo Ribeiro Nóbrega, Pedro Braga Neto, Danyela Martins Bezerra Soares, Fernando Regla Vargas, Clecio Godeiro, Paula Frassinetti Vasconcelos de Medeiros, Claudia Camejo, Maria Betania Pereira Toralles, Nelson Jurandi Rosa Fagundes, Laura Bannach Jardim, Maria Luiza Saraiva-Pereira Rede Neurogenetica

**Author notes:** **Corresponding author:** Laura Bannach Jardim, DMI FAMED UFRGS, and Medical Genetics Service, Serviço de Genética Médica, Hospital de Clinicas de Porto Alegre, Rua Ramiro Barcelos 2350, 90035-003 Porto Alegre, Brazil.

## Abstract

Spinocerebellar ataxia type 2 (SCA2) is an adult onset, dominant neurodegenerative disorder due to expansions of a CAG repeat tract at the *ATXN2* gene. A few studies on ancestral haplotypes were performed so far, and the allele C at rs695871 was always found in SCA2 carriers. We aimed to describe SCA2 ancestral haplotypes constructed based on the SNPs rs9300319, rs3809274, rs695871, rs1236900 and rs593226, using the STRs D12S1329, D12S1333, D12S1672 and D12S1332 to determine their genetic variation. Seventy-seven SCA2 families were recruited from Brazil, Peru, and Uruguay; 162 chromosomes from the Brazilian general population and the chromosomes with normal repeats from 101 SCA2 carriers were used as 263 controls. Eleven ancestral haplotypes were found in SCA2 families. The most frequent ones were A-G-C-C-C (46.7% of families), G-C-C-C-C (24.6%) and A-C-C-C-C (10.3%), with assigned risks of being associated with disease of δ = 0.326, 0.197 and 0, respectively. Their mean (sd) CAGexp were 41.68 (3.55), 40.42 (4.11) and 45.67 (9.70) (*p* = 0.055), while the mean (sd) CAG lengths at normal alleles were 23.85 (3.59), 22.97 (3.93) and 30.81 (4.27) (*p* < 0.001), respectively. The other SCA2 haplotypes were rare: among them, a G-C-G-A-T was found, evidencing a G allele in rs695871. In summary, our work identified eleven distinct SCA2 haplotypes in Brazilian, Uruguayan, and Peruvian families, including an unexpected SCA2 haplotype with a G allele at rs695871. These results suggest that SCA2 has multiple origins in these populations.

## 1. Introduction

The spinocerebellar ataxia type 2 (SCA2) is a neurodegenerative disorder caused by a dominant expansion of a (CAG)n tract located in the *ATXN2* gene that encodes for a protein named ataxin 2 (Pulst, 2015) ^1^. The upper limit for the length of normal alleles varies from 32 to 33 CAG repeats (Gardiner et al, 2019, de Castilhos et al, 2014) ^2,3^. Although the (CAG)n is polymorphic in normal chromosomes, the (CAG)_22_ allele is the most prevalent by far, reaching 90.1% of the general population (Andres et al, 2003; Akçimen et al, 2001) ^4, 5^.

The expanded CAG repeat (CAGexp) and/or the large polyglutamine (polyQ) tract in the coded protein, become toxic and lead to neurodegeneration primarily in the cerebellum and brainstem (Adegbuyiroa et al, 2017; Bunting et al, 2021) ^6, 7^. The main symptoms are progressive ataxia and dysarthria, slow saccadic eye movements, and peripheral neuropathy (Pulst, 2015) ^1^: their mean age of onset (AO) is 33.85 years and correlates with the CAGexp length (r²= 0.577; Sena et al, 2021) ^8^. CAGexp transmissions are unstable, with further expansions being the most frequent variation observed in the offspring (Almaguer-Medeiros et al, 2018) ^9^. As a consequence, anticipation is common and, in extreme cases, symptoms can start in childhood (Mao et al, 2002)^10^.

Anticipation also leads to a reduction in the reproductive period of CAGexp carriers. In fact, SCA2 individuals with early onset of symptoms have less children than those with late onset (Sena et al, 2019) ^11^. In this sense, anticipation would play a selective pressure against the maintenance of SCA2 in different populations.

Few haplotype studies have been carried out in SCA2 patients. Two reports reconstructed ancestral haplotypes exclusively with short tandem repeats (STRs). Based on D12S1672 and D12S1333, two SCA2 origins were determined in Japan (Mizushima et al, 1999) ^12^, while results based only on D12S1672 suggested a common origin in Cuban, English, Indian and Italian SCA2 families (Pang et al, 1999) ^13^. Three studies reconstructed haplotypes using the same single nucleotide polymorphisms (SNPs) rs695871 and rs695872, finding the C-C haplotype in all SCA2 families from Brazil, India, Italy, and Portugal (Choudhry et al, 2001; Ramos et al, 2010; Sonakar et al, 2021) ^14, 15, 16^. Although these results seem to contradict the hypothesis of multiple origins, since they proposed one common haplotype for SCA2 families living in distant geographical regions of the world, it is important to emphasize that the use of few markers makes the studies not very effective in detecting multiple origins of any given trait.

Haplotype reconstruction based on more markers and enhanced resolution is clearly needed to confirm or exclude the C-C haplotype as the proposed common ancestor of alleles causing SCA2. To help with this question, our study aimed to build ancestral SCA2 haplotypes using a larger number of markers in a wide sample of families from Brazil, Peru, and Uruguay.

## 2. Material and Methods

### 2.1 SCA2 carriers and controls

Subjects with a molecular diagnosis of SCA2 and living in Brazil, Peru and Uruguay between 2010 and 2022, were retrieved from the Rede Neurogenetica database (de Castilhos et al, 2014) ^3^, stored in protected files at the Hospital de Clinicas de Porto Alegre. Data about their families and their DNA samples were analyzed in this study. In addition, DNA samples from 81 subjects recruited from the general population of Rio Grande do Sul (RS) were obtained from an anonymous biorepository from our laboratory.

Informed consent was obtained from all individual participants. The study was approved by the Institutional Ethics Committee (Comissão de Ética em Pesquisa do Hospital de Clínicas de Porto Alegre) by the numbers CAAE 0324.1.001.000-06 and 02857818.6.0000.5327, GPPG 2006-0384, 2019-0169 and 2019-0254

### 2.2 Polymorphic markers

Five SNPs and four STR were used as polymorphic markers in this study, spanning a genomic region of 640 Kb (**Figure 1**). SNPs were selected if they had been previously used and if their MAF was of at least 0.35. SNP rs695871 has already been used in previous haplotype studies (Choudhry et al., 2001; Ramos et al., 2010; Sonakar et al., 2021) ^14, 15, 16^. SNPs rs9300319, rs3809274 and rs593226 were previously used in studies on positive selection of CAG repeats at *ATXN2* (Yu et al, 2005; Chen et al, 2013) ^17, 18^. Finally, rs12369009 was selected due to its disequilibrium binding with the CAG region. The rs695872 SNP was not genotyped since it is fully linked to rs695871 in more than 1200 chromosomes studied elsewhere (Choudhry et al, 2001; Ramos et al, 2010; Sonakar et al, 2021). ^14, 15, 16^

**Figure 1 -.**
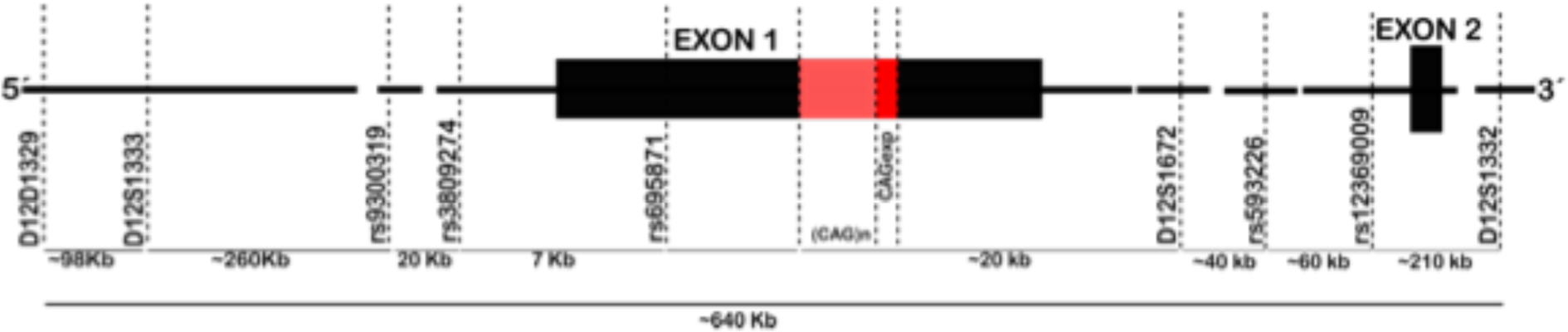
Molecular markers used for haplotype reconstruction in this study.

STRs D12S1329, D12S1333, D12S1672 and D12S11332 were used previously (Pang et al., 1999; Saleem et al., 2000; Choudhry et al., 2001; Ramos et al., 2010; Laffita-Mesa et al., 2012) ^13,19,14,15,20^, and were added here to build extended haplotypes.

### 2.3 Genotyping

DNA was isolated from peripheral blood leukocytes using standard methods. The CAG repeat length (CAGn) analysis was performed by the polymerase chain reaction (PCR) using fluorescent labeled primers flanking the CAG repeat tract at the *ATXN2* gene, followed by capillary electrophoresis into the 3500 Genetic Analyzer (Thermo Fisher Scientific). Results were analyzed through Microsatellite Analysis Software available at www.thermofisher.com.

The rs9300319, rs3809274, rs695871, rs12369009 and rs593226 SNPs genotyping were performed using TaqMan^®^ SNP Genotyping Assays (assays numbers C 1243837_10, C 1243827_10, C 7524190_30, C 30739839_10 and C 2978539_10, respectively) in a final volume of 8 µL containing 2 ng of DNA, according to the assay protocol (Thermo Fisher Scientific). Amplification was performed in the 7500 Fast Real-Time PCR System^®^ equipment (Thermo Fisher Scientific) as follows: one cycle of 50°C for 2 min, 95°C for 10 min, followed by 40 cycles of 95 °C for 15 s and 60 °C for 1 min.

Multiplex PCR was performed to identify the number of repeats at each STR. Amplifications were performed using 500 mmol/L dNTPs, 2.5 mmol/L MgCl_2_, 0.5 mmol/L DMSO, and 2 U of AmpliTaq Gold DNA polymerase (Thermo Fisher Scientific) in 1X AmpliTaq Gold buffer (Thermo Fisher Scientific), and 200 ng of genomic DNA. Four sets of fluorescently-labeled primers were used at 0.2 pmol: D12S1672 (5’ CAGGTGTGGTAGCACGA 3’; 5’ VIC-CTGGAAATTCACATCTGCTT 3’); D12S1333 (5’ FAM-TTCAGGTGGTACAGCCGT 3’; 5’ CATCAGAAGGCTTCATAGGAAT 3’); D12S1332 (5’ GCCAGGTACAGTGGCTC 3’; 5’ PET-CTGGGACCACAGGTGTAG 3’) and D12S1329 (5’ NED-CCTATCCCACCCAGGC 3’; 5’ AGTCTGCCCCAGGCAC 3’). The final volume of reaction was 25 uL. PCR cycling parameters were as follows: initial denaturation for 10 minutes at 94°C, followed by 30 cycles of 30 seconds at 94°C, 45 seconds at 60°C, and 90 seconds at 72°C, and a final extension at 72°C for 5 minutes. An aliquot (5 uL) of amplicon was mixed to Hi-Di^TM^ formamide (Thermo Fisher Scientific) and GeneScan™ 500 LIZ (Thermo Fisher Scientific) in a final volume of 10 uL, and capillary electrophoresis was performed in a 3530xL Genetic Analyzer (Thermo Fisher Scientific). Amplicon lengths were estimated using Microsatellite Analysis Software available at www.thermofisher.com.

### 2.4 Data analysis

Each SCA2 family was represented by one haplotype in the estimates of the relative frequencies of lineages. All chromosomes from subjects from the general population of South Brazil plus chromosomes carrying normal *ATXN2* alleles from up to one subject per Brazilian SCA2 sibship comprised the control chromosomes group, used to estimate the haplotypes present in the general Brazilian population. Brazilian SCA2 families were grouped according to their geopolitical regions of residence, as South, Southeast, Northeast, and North Brazil. The remaining SCA2 families were grouped as Peruvian and Uruguayan families.

Frequencies of SNP alleles obtained in SCA2 families and in the control chromosomes were compared using Fisher’s exact test with R Statistical Package.

Haplotypes were then inferred by segregation combined with the use of PHASE v2.1.1 (Stephens et al, 2001) ^21^. Reconstructed haplotypes with probabilities greater than 0.6 were used for analyses. Linkage disequilibrium analysis between normal and expanded alleles was tested by Fisher’s exact test. Evidence for LD was established using δ=(Fd-Fc)/(1-Fc), where Fd is the carrier frequency and Fc is the frequency of non-carrier chromosomes (Devlin and Risch, 1995; Gaspar et al, 2001)^22, 23^. When the haplotypic phase was not determined by the algorithm, chromosomes were not included in the analysis.

The haplotypes reconstructed with the five SNPs only (SNPs haplotypes) were considered the markers of SCA2 lineages and called ancestral haplotypes. The haplotypes reconstructed with the five SNPs plus the four STRs were called the extended haplotypes and were used to deduce the ancestral variants. The characteristics used to deduce that one extended haplotype was probably the ancestral of a given lineage, were (1) the frequency found in the expanded chromosomes, (2) the number of unchanged alleles, and (3) number of families with extended haplotypes just one step away from that putative ancestral haplotype (Martins et al, 2007) ^24^.

Associations between the resulting haplotypes and the number of CAG repeats in normal (*ATXN2*normal) and expanded (*ATXN2*exp) chromosomes were done using all the subjects studied.

Phylogenetic networks were performed through the Network 4.6.1.6 software (http://www.fluxus-technology.com) (Bandelt et al. 1999) ^25^, using microsatellite data. A combined reduced median and median-joining calculation was done to reduce reticulation. At first, phylogenetic networks were drawn using all nine molecular markers; later, STR markers were used to build phylogenies for SNP haplotypes, when possible. In all calculations, ɛ was set to zero, but different weights were attributed to the mutation rate in each marker. The ɛ value was adjusted for STRs, in an inversely proportional ratio to their variation in the length of the allele repeat (Martins et al, 2006)^26^.

To estimate genetic distances among SCA2 families, pairwise analyses were performed with Arlequin software version 3.5.2.2 (http://anthro.unige.ch/software/arlequin/software), using the sum of square size difference (R_ST_) as a measure of distance; R_ST_ being an analogue of F_ST_ suited for STR haplotype comparisons (Excoffier and Lischer, 2010) ^27^.

## 3. Results

One hundred and twenty individuals from 77 families carrying an expanded CAG repeat in *ATXN2* were included in this study. Sixty seven families were Brazilian, living in Rio Grande do Sul (22 families), São Paulo (29), Rio de Janeiro (5), Ceará (5), Rio Grande do Norte (2), Paraíba (1), Bahia (1), Pará (1), and Acre (1) states. Six Peruvian and four Uruguayan families were also included. Three out of 22 families from Rio Grande do Sul have been described previously (Ramos et al, 2010) ^15^. Samples from more than one related subject were obtained in 18 families (multiplex families).

A total of 241 chromosomes with normal *ATXN2* were used as population controls: 79 of them were the normal chromosomes from SCA2 carriers, and 162 were the chromosomes from 81 subjects recruited from the population of Rio Grande do Sul state.

### 3.1 SNPs and ancestral haplotypes

Allelic frequencies of rs9300319, rs3809274, rs695871, rs12369009 and rs593226 in the 77 expanded *ATXN2* alleles and in the 263 normal *ATXN2* alleles were described in **Table S1.** The SNPs frequencies found in the expanded *ATXN2* alleles (*ATXN2*exp) were significantly different from those found in the normal *ATXN2* alleles (*ATXN2*normal), with the exception of rs3809274 (**Table S1**). The median (range; variance) number of CAG repeats associated with C and G alleles in normal chromosomes were 24.40(14-33; 24.40) and 22.43 (21-33; 3.36); the (CAG)_22_ was found in 42.55% of C and in 89.45% of G alleles.

The 24 haplotypes found in *ATXN2*exp and *ATXN2*normal alleles are shown in **Table 1**, as well as their frequencies. The geographic distribution of the eleven ancestral haplotypes found in SCA2 families is presented in **Figure 2**. Three ancestral haplotypes were frequently found in SCA2 carriers and in controls. The A-G-C-C-C haplotype was present in SCA2 families from all geographic regions, being the most frequent (46.7%) among them. The attributed risk of A-G-C-C-C being associated with SCA2 was δ = 0.326. G-C-C-C-C haplotype was present in controls and in SCA2 families from Brazil, but not from Peru or Uruguay. G-C-C-C-C had an assigned risk of being associated with SCA2 of δ = 0.263. A-C-C-C-C was found in 10.3% of the SCA2 families; its frequency among controls was similar and was unrelated to an increased risk of SCA2. **Table 2** presents the distribution of the CAGn in *cis* with these three ancestral haplotypes, in normal and in expanded alleles.

**Figure 2 -.**
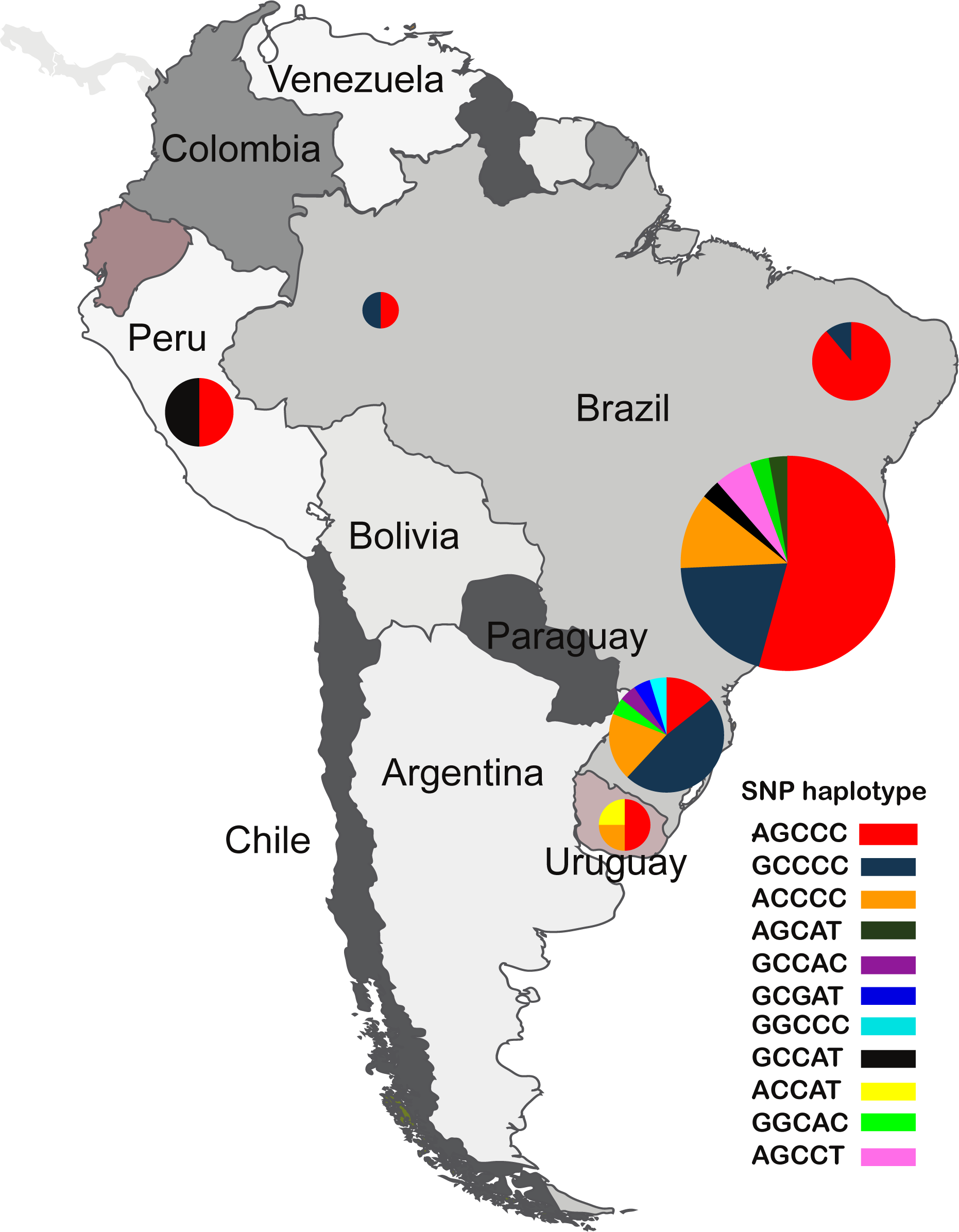
Geographic distribution of SNP haplotypes in SCA2 families.

**Table 1 -.**
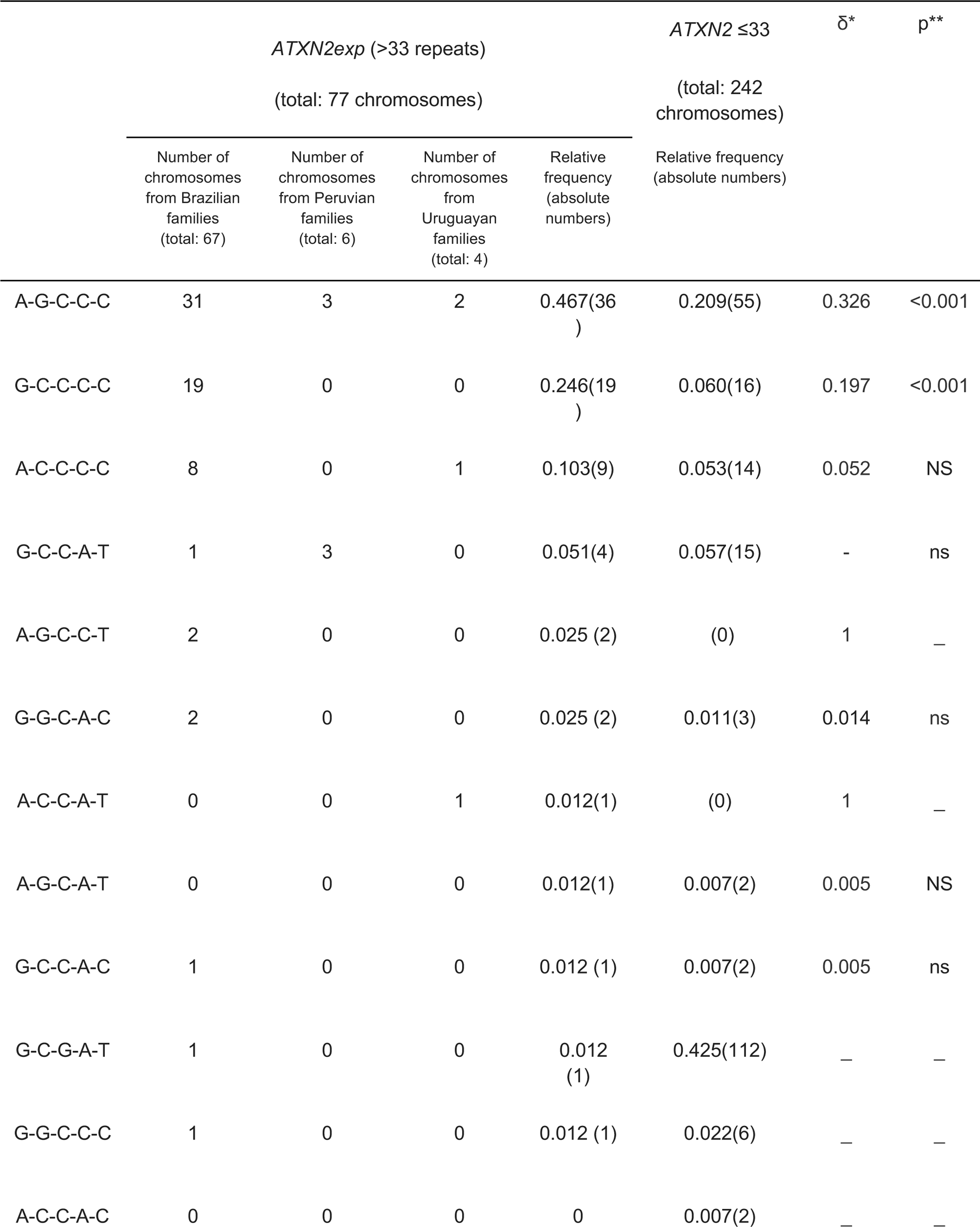
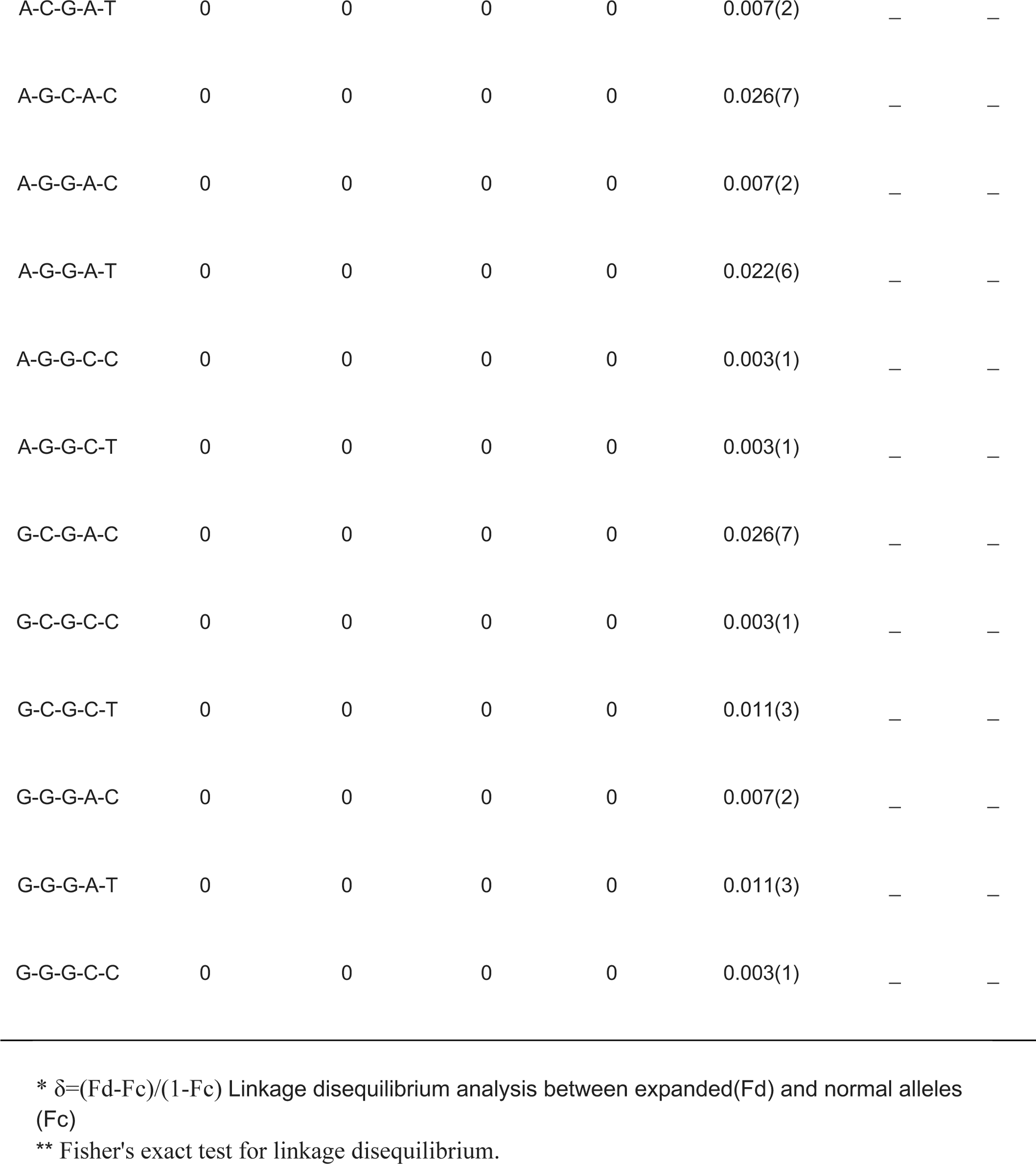
Frequency of SNP haplotypes in SCA2 families and control chromosomes.

**Table 2.** The mean CAG of the most frequently expanded and normal SNP haplotype.

|  | SNP haplotype | n<br>(origin)** | mean CAG(SD) | p * |
| --- | --- | --- | --- | --- |
| Expanded CAG | A-G-C-C-C | 36 | 41.68 (3.55) | 0.055 |
|  | G-C-C-C-C | 19 | 40.42 (4.11) |  |
|  | A-C-C-C-C | 9 | 45.67 (9.70) |  |
| Normal CAG | A-G-C-C-C | 55<br>(36, 18 and 1 chromosomes from<br>South, Southeast and Northeast<br>Brazil, respectively) | 23.85 (3.59) | < 0.001 |
|  | G-C-G-A-T | 112<br>(83, 17, 4 and 1 chromosomes from<br>South, Southeast, Northeast and<br>North Brazil, and 4 and 3<br>chromosomes from Peru and<br>Uruguay, respectively) | 22.57 (2.04) |  |
|  | G-C-C-C-C | 16<br>(12 and 4 chromosomes from<br>South and Southeast Brazil,<br>respectively) | 30.81 (4.27) |  |
|  | A-C-C-C-C | 14<br>(9, 3 and 1 chromosomes from<br>South, Southeast and Northeast<br>Brazil, respectively, and 1<br>chromosome from Uruguay) | 22.97 (3.93) |  |

\* Kruskal-Wallis test
\*\* for the control group, only. Please check Table 1 for SCA2 carriers.

Other six ancestral haplotypes were found in controls and in few SCA2 families each, so that further analyses were not granted. One of them deserves special attention, since it is the first report of the presence of a G allele in rs695871 in SCA2 carriers: the haplotype G-C-G-A-T, found in three individuals from one SCA2 family of mixed ancestry from Southern Brazil (**Figure 2**, in navy blue, and **Table 1**).

Finally, two ancestral haplotypes were found in SCA2 families but not in controls: A-C-C-A-T and A-G-C-C-T (**Table 1**).

### 3.2 STRs and extended haplotypes

When all SNPs and STRs were used as markers, 187 distinct extended haplotypes were found in cases and controls. Thirty-one extended haplotypes segregated within SCA2 families (**Table S2**), while 160 were present in the control group only (**Table S3**). SCA2 families and controls shared four extended haplotypes as follows: 17-17-G-C-G-21-A-T-23, 17-19-G-C-C-21-C-C-23, 17-19-G-G-C-21-C-C-23, and 18-12-A-G-C-22-C-C-26. The former three were considered the ancestors of their lineages.

The CAGn at *ATXN2* of the control individuals that shared the four extended haplotypes with SCA2 carriers are presented in **Table 3**. All control subjects carrying the extended haplotypes considered the SCA2 lineage ancestors 17-19-G-C-C-21-C-C-23 and 17-19-G-G-C-21-C-C-23 had 33 CAG repeats, and two out of 16 controls with 17-17-G-C-G-21-A-T-23 had 31 CAG repeats in *cis.* It is also worthy of note to mention that the CAGn variances of control individuals who shared and share not their extended haplotypes with the SCA2 families were of 28.22 and 7.36, respectively (**Table 3**).

**Table 3 -.** CAG repeat lengths related to the extended haplotypes found in the control group.

|  | Haplotype | CAG repeat<br>length<br>mean<br>(variance) | Individual data |  |
| --- | --- | --- | --- | --- |
|  |  |  | CAG repeat<br>lengths<br>found | number of<br>carriers |
| Shared by<br>controls and<br>SCA2 families | 17-17-G-C-G-21-A-T-23 | 23.12(9.45) | 22 | 14 |
|  |  |  | 31 | 2 |
|  | 17-19-G-C-C-21-C-C-23 | 33(0.00) | 33 | 7 |
|  | 17-19-G-G-C-21-C-C-23 | 33(0.00) | 33 | 2 |
|  | 18-12-A-G-C-22-C-C-26 | 22(0.00) | 22 | 1 |
|  | all shared haplotypes | 26.5 (28.22) |  |  |
| Only found in<br>controls | All the others | 22.87(7.36) |  |  |

The extended haplotypes found in more than one SCA2 family were 26-12-A-G-C-23-C-C-23, 17-19-G-C-C-21-C-C-23, 17-13-A-G-C-23-C-C-25 and 18-16-A-C-C-17-C-C-23, shared by 18, 14, 4 and 4 families, respectively. Of these, 17-19-G-C-C-21-C-C-23 was the only one found in the control group, where it was related to normal large CAG repeats (**Tables S2 and S3**).

The haplotype trees of the common lineages A-G-C-C-C, G-C-C-C-C and A-C-C-C-C were further detailed in **Figure 3**. A-G-C-C-C was detected in SCA2 families from all geographic regions under study, and presented a genetic diversity of 0.7397 (+/− 0.074) and average gene diversity over loci of 0.491270 (+/− 0.316391). The most conservative interpretation suggested that 26-12-A-G-C-23-C-C-23 was the founder or the older ancestor of the lineage A-G-C-C-C, since this extended haplotype was the most frequent among SCA2 families, and three families might have derived from it with a distance of one mutational step (**Figure 3A**). G-C-C-C-C (**Figure 3B**) has the lowest genetic diversity - 0.5158 (+/− 0.1316) - and average gene diversity over *loci* - 0.405263 (+/− 0.279203). The extended haplotype 17-19-G-C-C-21-C-C-23 was defined as the ancestor; curiously, it was also shared by SCA2 and normal chromosomes. Finally, the lineage A-C-C-C-C (**Figure 3C**) presented the greatest genetic diversity 0.7778 (+/− 0.110) and average gene diversity over *loci*: 0.486111 (+/− 0.344187).

**Figure 3 -.**
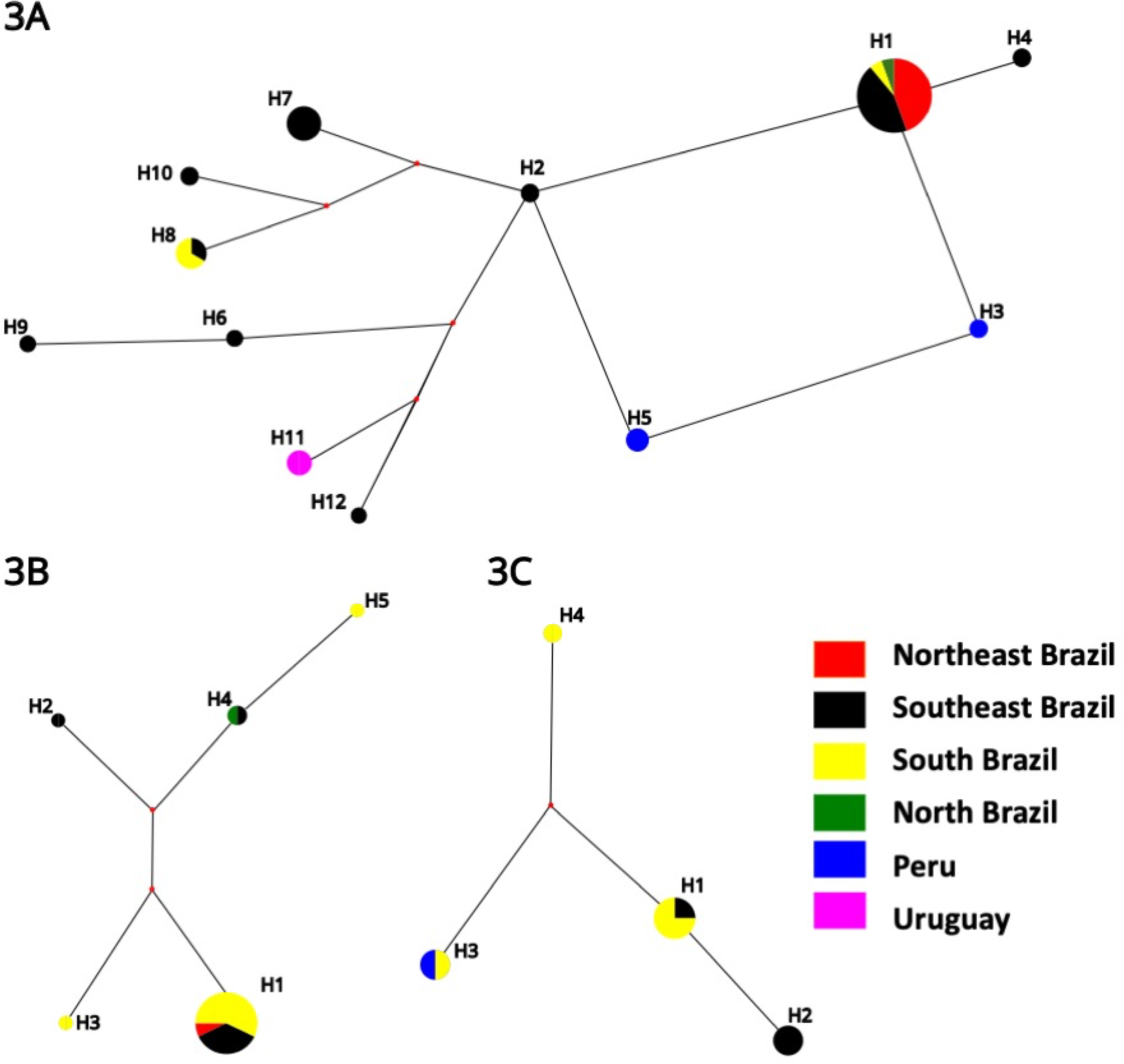
Phylogenetic networks of the three haplotypes based on 4 microsatellites. Circles and line sizes are proportional to number of families and stepwise mutation, respectively. The color in circle: South, southeast, northeast, and north of Brazil in the colors yellow, black, red and green respectively, Peru blue and Uruguay brown, represent the region where haplotypes were found. **(A)** A-G-C-C-C **(B)** G-C-C-C-C **(C)** A-C-C-C-C.

## 4. Discussion

Eleven different ancestral *ATXN2*exp lineages were identified in our South American cohort: A-G-C-C-C (46.7% of *ATXN2*exp families), G-C-C-C-C (24.6%), A-C-C-C-C (10.3%), G-C-C-A-T (5.1%), G-G-C-A-C (2.5%), A-G-C-C-T (2.5%), and A-C-C-A-T, A-G-C-A-T, G-C-C-A-C, G-C-G-A-T, and G-G-C-C-C (each one of the last five combinations present in one or 1.2% of *ATXN2*exp families). A-G-C-C-T and A-C-C-A-T were missing from our control group; if they were derivatives from one of the former haplotypes, such as A-G-C-A-T, remains to be established. The discovery of a Brazilian SCA2 family carrying the G-C-G-A-T haplotype refuted the current hypothesis that expansions would be universally linked to C allele in rs695871. In general terms, these results denoted that the *ATXN2*exp have multiple origins, and may, rarely, occur in the most protected *ATXN2* background linked to the G allele in the rs695871.

Before our study, the available data on the ancestral origins, population frequencies and genetic characteristics of the *ATXN2*norm and *ATXN2*exp alleles formed a heterogeneous and intriguingly contradictory set. On one hand, SCA2 was characterized by very intense anticipations (Filla et al, 1999; Sena et al, 2019; Gamberdella et al, 1998; Burk, 1997) ^28,11, 29, 30^, suggesting that the lineages would have rapid extinctions (Sena et al, 2021) ^8^. On the other hand, all previous studies that used standardized markers to reconstruct haplotypes found a single ancestral pattern, the C-C at rs695871 and rs695872 (Choudhry et al, 2001; Ramos et al, 2010; Sonakar et al, 2021) ^14, 15, 16^, in families living worldwide. The G allele in rs695871 was associated to 70.9% of Indian (Choudhry et al, 2001) ^14^ and to 89.45% of the present Brazilian controls carrying the (CAG_22_) allele. Considering that (CAG_22_) is very stable, descendent chromosomes would have a negligible chance of ever expanding and giving rise to a new SCA2 lineage. In contrast, the variable CAGn related to C-C pattern (or simply C at rs695871) - 77.4% of them different from the (CAG_22_) - would allow for subsequent instabilities and for expansions to reach the range associated with the disease. The data so far obtained “have provided the support for a limited pool of ‘ancestral’ or ‘at risk’ haplotypes from which SCA2 disease chromosomes are derived” (Choudhry et al, 2001) ^14^.

Despite these data, a large number of different ancestral haplotypes was always a reasonable alternative for SCA2, in view of the severity of anticipation phenomena known to occur in this disease.

A mathematical model that combines anticipation, fitness and segregation distortion estimated that the median (range) survival of an expanded allele after its *de novo* appearance would be 10 (1 to 121) generations, or 250 (25 to 3,000) years (Sena et al 2023) ^31^. Given that SCA2 populations were found worldwide, this short estimated survival of lineages led to the proposal that SCA2 lineages should have multiple origins, related or not to haplotypes predisposed to expansions. Descriptions of *de novo* expansion carriers would favor this hypothesis. However, they are quite rare, such as one individual with 22/35 CAG repeats at *ATXN2* with asymptomatic mother and father carrying 22/22 and 22/32 repeats (Futamura et al, 1998) ^32^.

Our findings provide further evidence in accordance with these expectations, since eleven ancestral lineages were discovered in our South American cohort. At this point it was not possible to clarify the genetic distances and relationships between all lineages, even if some of them presented similitudes - for instance, if A-G-C-C-T could be a derivative of A-G-C-A-T, or vice-versa. Neither our markers nor the population under study allow any speculation in this direction. That said, we will move on to discuss the results of the most informative ancestral haplotypes found in our study.

A-G-C-C-C was the most frequent haplotype found, being present in 36 (46.7%) SCA2 families living in all regions studied. Since A-G-C-C-C was present in 5.3% of control chromosomes, it was related to a relatively high risk for carriers of having an *ATXN2*exp (the δ of 0.467). 26-12-A-G-C-23-C-C-23 was the most frequent extended haplotype; being so common in our cohort, A-G-C-C-C might well be the most common elsewhere. A global study including SCA2 families from Europe, Africa and Asia is needed to clarify the A-G-C-C-C origin.

G-C-C-C-C was the second most common ancestral haplotype found in our SCA2 families, with a widespread geographical distribution across Brazil and Peru. Several genetic characteristics made us think that this haplotype might be also associated with an unstable repeat, prone to expansions and consequently to *de novo* mutations originating from an ancestral *ATXN2*normal haplotype.

G-C-C-C-C was only found in Brazilian SCA2 carriers and had the lowest genetic diversity in SCA2 families, suggesting a common and recent origin. Moreover, the mean (SD) CAGn in unexpanded G-C-C-C-C was larger than the length found in the remaining normal chromosomes (**Table 2**); actually, the seven control chromosomes that carried the extended haplotype 17-19-G-C-C-21-C-C-23 had 33 CAG repeats (**Table 3**), a CAG length that in other populations has been related to SCA2 with very late onset (Santos et al, 1999; Futamura et al, 1998; Fernandez et al, 2000) ^32, 33, 34^. In contrast, SCA2 individuals with this extended haplotype 17-19-G-C-C-21-C-C-23 carried a mean (SD) of 39.53 (3.65) CAGn, while other SCA2 subjects) carried 42.50 (5.18) repeats (p = 0.0123). These findings suggest that G-C-C-C-C expanded repeats might have recently arisen from a normal G-C-C-C-C reservoir linked to normal, long *ATXN2* alleles; probably 17-19-G-C-C-21-C-C-23. We cannot be sure of that, since most of our controls originated from South Brazil (**Table 2**). Similar phenomena were seen in spinocerebellar ataxia type 3, also known as Machado-Joseph disease (SCA3/MJD), where the average length of CAGexp was longer in the A-C-A than in the G-G-T haplotypes, and where larger CAGexp were associated with older lineages and smaller CAGexp, with younger lineages (Martins at al, 2008)^35^. However, there are some facts hard to be connected with a common and recent origin for all G-C-C-C-C SCA2 carriers from our study: the very large geographical distances between families, ten of them living in South, seven in Southeast, one in Northeast and one in North regions of Brazil (**Figure 2**); and the lack of some connecting branches between extended haplotypes of SCA2 carriers in the phylogenetic tree (**Figure 3B**).

Although this hypothesis remains to be better studied, G-C-C-C-C has some characteristics of a haplotype potentially prone to undergo subsequent expansions towards SCA2. The hypothesis of a predisposing haplotype was raised in early studies on C-C haplotype in *ATXN2* (Choudhry et al, 2001) ^14^. A similar phenomenon was described in the *HTT* gene, where the A1 and A2 haplotypes were associated with longer normal CAGn and with a greater risk of generating *de novo* mutations in Huntington’s disease (HD) (Warby et al, 2011) ^36^. Although plausible, this assumption needs to be confirmed by studying a larger number of *ATXN2* controls from different populational origins.

A-C-C-C-C was found in nine SCA2 families, located in South and Southeast Brazil, as well as in Uruguay. There are no extended A-C-C-C-C haplotypes in common between controls and SCA2 families. This might suggest that this lineage entered the local populations recently - for instance, from a migration originating from the Iberic peninsula or from Italy, both being reasonable alternatives based on the history of European occupation of these Latin American regions. The CAGexp length at A-C-C-C-C was larger, though non-significant, than other *ATXN2*exp, and we speculate that A-C-C-C-C expansion followed a mechanism distinct from G-C-C-C-C, as A-C-C-C-C was associated with 22.92 ± 3.93 repeats in normal subjects (**Table 2**).

Among the rare SCA2 haplotypes found here, G-C-G-A-T stood out for the unexpected finding of a G allele in rs695871. As mentioned before, previous published data found that a common haplotype (C^rs695871–^C^rs695872^) was present in 100% of Indian, Portuguese, Italian and three Brazilian families also included in this study (Choudhry et al, 2001; Ramos et al, 2010; Sonakar et al, 2021) ^14, 15, 16^. Our G-C-G-A-T family greatly increases the variety of origins of this condition, by identifying a lineage not even outlined.

Nevertheless, we are aware that our study has some limitations. The classic marker in rs695872 was not included in the study. However, it is unlikely that the data obtained with this marker would add new information, as the alleles in rs695871 and rs695872 are 100% linked - i.e., C at rs695871 is always linked to C at rs695872. In addition, CAGexp internal interruptions, in turn, could be very informative for the reconstruction of haplotypes, and this was not addressed by our study.

On the other hand, the history of several bottlenecks to which Latin American populations were exposed in the last and recent 500 years prevented us from performing reliable dating of the detected lineages. There was the Amerindian genocide - bigger in Brazil and Uruguay than perhaps in Peru, countries of origin of SCA2 families included in this study. At the same time, the European occupation was carried out in several relevant migratory waves and even depended on the forced migration of a huge number of enslaved Africans. The last major migrations took place between 1825 and 1905, with the arrival of non-Iberian Europeans mainly from Italy, Germany, Poland, and Ukraine. Numerous founder effects have been proposed for geographic isolates on the continent, including that of SCA2 in Holguin, Cuba (Rodriguez-Labrada et al, 2020) ^37^. It is plausible to imagine that many of the lineages described here come from recent migrations. It is also possible, but very speculative, to propose an Amerindian origin to G-C-G-A-T. If so many ancestral haplotypes occur in South American families, it is easy to to predict the existence of more lineages among other ethnic groups. The heterogeneity of the South American population contributed to confirm the existence of several origins, by including recent Amerindians, European and African ancestries. However, the South American population structure changed significantly in recent centuries. The study on the origins of SCA2 should have a global scale to allow more consistent datation of lineages.

In conclusion, this work identified eleven distinct ancestral haplotypes in Brazilian, Uruguayan, and Peruvian SCA2 families, suggesting multiple origins for SCA2 in these populations. The existence of multiple origins indicates that *de novo* mutations might play an important role in the maintenance of SCA2 in the population. A global, multicentric study is needed to infer the number of origins of SCA2 and their respective dating, as well as to detect other haplotypes associated with expanded *ATXN2*.

## Data Availability

All data produced in the present study are available upon reasonable request to the authors

## 6. Acknowledgements

The authors are grateful to the individuals who agreed to participate in this study.

This study was supported by Fundo de Incentivo à Pesquisa do Hospital de Clínicas de Porto Alegre (FIPE-HCPA), grants number GPPG 2006-0384, 2019-0169 and 2019-0254; and DNABank Neurogenetics-INCH Lima-Peru trough grant number ASAP/GP2 - MJFF-023323. LSS, MLSP and LBJ were supported by CNPq.

## 7. Authors contribution

L.S.S., G.V.F., M.L.S.P., and L.B.J. contributed to the conception and design of the study; L.S.S., G.V.F., J.L.P., O.B., M.C.O., P.R.N., P.B.N., D.M.S., F.R.V., C.G., P.F.V.M., C.C., M.L.S.P., L.B.J. contributed to the acquisition of data; L.S.S., G.V.F., and L.B.J. analyzed the data; L.S.S. and L.B.J drafted the text; L.S.S. prepared the figures. All authors reviewed the manuscript.

## 1. Supplemental Data

**Table S1.** Allelic frequencies of rs9300319, rs3809274, rs695871, rs12369009, and rs593226 in SCA2 families and controls included in this study.

**Table S2.** Extended haplotypes found in SCA2 and in normal chromosomes found in the present study

**Table S3.** All the extended haplotypes found in the present study

